# A data driven approach to address missing data in the 1970 British birth cohort

**DOI:** 10.1101/2024.02.01.24302101

**Authors:** Michail Katsoulis, Martina Narayanan, Brian Dodgeon, George Ploubidis, Richard Silverwood

**Affiliations:** MRC unit for Lifelong Health and Ageing, Institute of Cardiovascular Science, UCL; Centre for Longitudinal Studies, Institute of Education, UCL

## Abstract

**Background:** Missing data may induce bias when analysing longitudinal population surveys. We aimed to tackle this problem in the 1970 British Cohort Study (BCS70)

**Methods:** We utilised a data-driven approach to address missing data issues in BCS70. Our method consisted of a 3-step process to identify important predictors of non-response from a pool of ∼20,000 variables from 9 sweeps in 18037 individuals. We used parametric regression models to identify a moderate set of variables (predictors of non-response) that can be used as auxiliary variables in principled methods of missing data handling to restore baseline sample representativeness.

**Results:** Individuals from disadvantaged socio-economic backgrounds, increased number of older siblings, non-response at previous sweeps and ethnic minority background were consistently associated with non-response in BCS70 at both early (ages 5-16) and later sweeps (ages 26-46). Country of birth, parents not being married and higher father’s age at completion of education were additional consistent predictors of non-response only at early sweeps. Moreover, being male, greater number of household moves, low cognitive ability, and non-participation in the UK 1997 elections were additional consistent predictors of non-response only at later sweeps. Using this information, we were able to restore sample representativeness, as we could replicate the original sample distribution of father’s social class and cognitive ability and reduce the bias due to missing data in the relationship between father’s socioeconomic status and mortality.

**Conclusions:** We provide a set of variables that researchers can utilise as auxiliary variables to address missing data issues in BCS70 and restore sample representativeness.

**Key Messages:**

- We aimed to address the problem of missing data in the 1970 British Cohort Study (BCS70) caused by non-response at different sweeps
- We identified a set of predictors of non-response that can successfully restore baseline sample representativeness across sweeps
- The information from this study can be used from researchers in the future to utilise appropriate auxiliary variables to tackle problems due to missing data in BCS70

## INTRODUCTION

Missing data are a significant limitation when analysing longitudinal population surveys[1]. Researchers in social and biomedical sciences aim to address specific research questions related to: i) describing certain characteristics in the population at specific time points, ii) developing prediction algorithms to model the outcome of interest, and iii) estimating the causal effect of an exposure on an outcome[2]. In all of these cases, missing data can greatly impact the analysis and introduce bias[3-4]. Nearly half a century ago, Rubin introduced a framework to characterise missing values[5]. This framework categorizes missingness mechanisms into three broad classes: a) missing completely at random (MCAR), where the probability of missing data does not depend on observed or unobserved variables, or in the context of a longitudinal population survey that missingness is not related to any characteristics of the participants, b) missing at random (MAR), where the probability of missing data depends only on observed variables, or observed characteristics of participants, and c) missing not at random (MNAR), where the probability of missing data depends on unobserved variables, or unobserved characteristics of the participants. Complete-case analysis is the simplest approach to handle missing data, but it is only valid when the missing data mechanism is MCAR and in certain situations for MAR and MNAR[6]. Moreover, complete-case analysis may have limited statistical power, depending on the proportion of observations with missing data. Therefore, for MAR, more sophisticated approaches such as inverse probability weighting (IPW)[7], full information maximum likelihood (FIML)[8] and multiple imputation (MI)[3-4] should be considered, whereas when dealing with MNAR, further assumptions are needed to appropriately account for the missingness mechanism[9].

In the analysis of longitudinal surveys with missing data, it is common for the missing data mechanism to be either MAR or MNAR[10]. To enhance the plausibility of the MAR approach, it is recommended to consider auxiliary variables along with the variables of substantive interest, particularly when these variables are also associated with the variable(s) of interest affected by missingness[11].

In this paper, our objective is to implement a systematic and principled approach to address the challenges posed by missing data in the 1970 British Cohort Study (BCS70)[12-13]. We leverage the rich and comprehensive information from over 18,000 participants across 10 survey sweeps in BCS70. Our approach utilises auxiliary variables to enhance the robustness of analyses subject to missing data. We demonstrate how our methodology can be applied to strengthen the plausibility of the MAR assumption and restore sample representativeness, thereby improving the validity of the subsequent analysis.

## METHODS

### Data

BCS70 is an ongoing multidisciplinary birth cohort study consisting of 18,000 participants born in 1970, with 10 major sweeps including at birth (sweeps 0 to 9). The initial sample has been supplemented with migrants at ages 5, 10 and 16 and the most recent completed follow-up was at age 46 (in 2016), with data currently being collected as part of the age 51-53 (2021-2023) sweep[13]. This cohort contains high quality prospective data on health, physical, educational and social development, and economic circumstances among other factors documented in >20,000 variables across sweeps.

We additionally utilised the Health Survey for England (HSE) to derive hypothetical external benchmark estimates of body mass index (BMI) at age 34[14].

### Predictors of non-response

BCS70 datasets from all sweeps up to sweep 8 (age 42) contain variables that could potentially be considered as predictors of non-response at subsequent sweeps. We aimed to identify the important predictors of non-response in BCS70, using a similar process to that applied in NCDS[1]. Specifically, we excluded “routed” variables (questions that depend on a specific response to a previous question), used index/score variables that combined information across variables rather than the constituent items where applicable, excluded all binary variables with prevalence <1%, categorical variables with >98% in one category and variables with >40% of data missing, and recoded categorical variables with <1% in one category when possible. After cleaning all BCS70 waves according to these guidelines, 967 variables remained as eligible predictors of non-response.

### Definition of non-response

We created a dichotomous variable (yes/no) to define non-response in each sweep from sweep 1 (age 5) onwards. A participant was considered a non-respondent if they did not take part in the survey at the specific sweep, either due to refusal, or because establishing contact was not possible or was not attempted (e.g. because of long-term refusal) (see Table 1). We excluded participants who died or emigrated from our analysis because our primary focus was to identify predictors of non-response, rather than examining mortality or emigration. Our approach to handling missing data aims at restoring the representativeness of our sample relative to a well-defined target population. For BCS70, the target population at each sweep consists of all participants born in 1970 who are alive and residing in Great Britain at that specific age.

**Table 1.** Participation in the 1970 British cohort study from birth to 46 years.

|  | Total cohort | Dead | Emigrants | Eligible sample | Respondents, n (%) | Non-Respondents, n (%) |
| --- | --- | --- | --- | --- | --- | --- |
| Sweep 0 - Birth | 18037* | 0 | 0 | 18037 | 16589 (92%) | 1448 (8%) |
| Sweep 1 - Age 6 | 18037* | 569 | 0 | 17468 | 13135 (75%) | 4333 (25%) |
| Sweep 2 - Age 10 | 18037* | 590 | 0 | 17447 | 14867 (85%) | 2580 (15%) |
| Sweep 3 - Age 16 | 18037* | 623 | 0 | 17414 | 11615 (67%) | 5799 (33%) |
| Sweep 4 - Age 26 | 18037 | 716 | 34 | 17287 | 9003 (52%) | 8284 (48%) |
| Sweep 5 - Age 30 | 18037 | 767 | 235 | 17035 | 11261 (66%) | 5774 (34%) |
| Sweep 6 - Age 34 | 18037 | 820 | 432 | 16785 | 9665 (58%) | 7120 (42%) |
| Sweep 7 - Age 38 | 18037 | 882 | 456 | 16699 | 8874 (53%) | 7825 (47%) |
| Sweep 8 - Age 42 | 18037 | 966 | 433 | 16638 | 9841 (59%) | 6797 (41%) |
| Sweep 9 - Age 46 | 18037 | 986 | 466 | 16585 | 8591 (52%) | 7994 (48%) |

\*Individuals who were not resident in the UK in 1970 (here included as not respondents) were added later at sweep 1-3. For more details, see
https://doc.ukdataservice.ac.uk/doc/5641/mrdoc/pdf/bcs70_responses_and_deaths_user_guide.pdf

### Analytic strategy

We followed a 3-stage analytic approach using the 967 eligible variables as input, similar to the strategy employed in NCDS[1]. At each stage we modelled non-response with a Poisson model with robust standard errors (modified Poisson regression[15]) that returns risk ratios to avoid bias due to the non-collapsibility of the odds ratio[16] as non-response after age 16 becomes more common (>30%). Below, we describe the 3-stage procedure for non-response at sweep t:

Stage 1: Univariable modified Poisson regressions of non-response at sweep t on each individual potential predictor of non-response at sweep 0 up to sweep t –1 in complete-case analysis. Keep predictors with p-value<0.001.

Stage 2: Multivariable modified Poisson regressions of non-response at sweep t on all predictors retained from stage 1, separately from sweep 0 up to sweep t –1, in complete-case analysis. Keep predictors with p-value<0.05.

Stage 3: MI using all retained variables plus non-response at sweep t in the imputation model. MI multivariable modified Poisson regressions for all retained predictors at sweep 0, up to sweep t –1, adjusted for predictors at all previous (but not subsequent) sweeps. Keep predictors with p-value <0.001.

Sex, country of birth, participation in all previous sweeps and father’s socioeconomic status were omitted from stages 1 and 2 and instead included a priori in Stage 3. The detailed procedure is described in the Appendix, Section 1.

### Restoring sample representativeness

We explored whether the identified predictors of non-response at sweep 9 (age 46) had the potential to restore sample representativeness with respect to paternal social class at birth and cognitive ability assessed at age 5 (estimated by the first principal component of the variables “copying designs test score”, “English picture vocabulary test”, “human figure drawing test” and “complete-a-profile test score”). We compared the following estimates of percentage in each social class/mean cognitive ability: i) using all available data at birth/age 5 (forming the “known truth” comparator); ii) using data only from respondents at age 46 (i.e., discarding data from non-respondents at age 46) to assess the extent of non-response bias; and iii) using MI including the identified predictors of non-response at sweep 9 as auxiliary variables, to see whether sample representativeness (relative to the previously estimated “known truth”) could be restored.

Additionally, we estimated the hazard ratios of adult mortality (age>26 years) by categories of paternal social class at birth using Cox regression. We then discarded non-respondents at sweep 4 (age 26) and looked at this relationship again with a complete-case analysis. Finally, we estimated the association of paternal social class and adult mortality, using MI, including the identified predictors of non-response at sweep 4 as auxiliary variables in the imputation models.

We also present how external validation can be utilised in BCS70 in the Appendix (Section 2). Our example was based on the estimation of mean BMI levels for men and women at age 34 (wave 6), using data from HSE in 2004 as a hypothetical example of an external benchmark.

## RESULTS

### Non-response in BCS70

Table 1 provides descriptive statistics on participation in BCS70 from birth to age 46 years. As anticipated, participation rates decrease over time, with notable declines observed at specific ages. Non-response can be considered in two different periods; before and after age 16. The reason is the interviews in the first 4 sweeps (sweep 0 to 3) were conducted mainly with the parents (up to age 16), while afterwards the interviews were performed with the cohort members themselves (in age 16 both parents and participants were interviewed)[18]. The participation rate with respect to the eligible sample of each sweep is between 67%-85% up to sweep 3, while the participation rate ranges from 52-66% in the following sweeps. Among the total 18,037 cohort members[19], 3183 (17.6%) took part in all 10 sweeps and 986 (5.5%) died before age 46 (sweep 9). Of note, as in Table 1, of the 18037 cohort members, 16589 participated at the birth sweep, while the remaining cohort members were immigrants that were added to the sample in later sweeps. Among the 16,585 eligible (alive and not emigrated) cohort members at age 46, 7994 (48.2%) were non-respondents.

### Predictors of non-response in BCS70

In the Appendix (Section 1), we provide detailed insights into our process for identifying predictors of non-response across all sweeps of BCS70. The number of predictors of non-response ranges from 7 (for non-response at sweep 1) to 16 (for non-response at sweep 9). Additionally, in Tables 2 and 3, we present the results for the “consistent” predictors of non-response observed in sweeps 1-3 (involving interviews with parents) and sweeps 4-9 (involving interviews with participants who have reached adulthood). We define “consistent” predictors as those variables that were identified for more than half of the subsequent sweeps, either in sweeps 1-3 or sweeps 4-9. These predictors demonstrate a recurring relationship with non-response across multiple survey waves, suggesting their potential relevance in understanding and addressing missing data. In sweeps 1-3, the consistent predictors of non-response were cohort member’s (or their parent’s) participation in all previous sweeps, country of birth, father’s disadvantaged socioeconomic status, parents not being married, increased father’s age at completion of education, increased number of older siblings and method of contraception (Table 2). Cohort members’ participation, father’s socioeconomic status and number of older siblings continued to be predictors of non-response in these later sweeps (Table 3). However, we also identified different predictors of non-response in this stage. Specifically, being male, household moves, cognitive ability at age 5 and 10, and (non-) voting in the election in 1997 emerged as consistent predictors of non-response.

**Table 2:** Estimated risk ratios and 95% confidence intervals for consistent predictors (selected in at least 50% of possible sweeps) of non-response at sweeps 1-3 (ages 5-16) in the 1970 British Cohort Study.

|  | Sweep 1 (age 5) |  | Sweep 2 (age 10) |  | Sweep 3 (age 16) |  |
| --- | --- | --- | --- | --- | --- | --- |
|  | RR | 95% CI | RR | 95% CI | RR | 95% CI |
| **Non-response at previous sweeps** |  |  |  |  |  |  |
| Incomplete vs complete response | NA | NA | 4.53 | 4.20, 4.89 | 1.54 | 1.43, 1.66 |
| **PREDICTORS from SWEEP 0 (age 0)** |  |  |  |  |  |  |
| **Country of birth** |  |  |  |  |  |  |
| England | Ref |  | Ref |  |  |  |
| Wales | 0.69 | 0.59, 0.81 | 0.65 | 0.53, 0.80 | 0.70 | 0.62, 0.79 |
| Scotland | 1.13 | 1.04, 1.23 | 0.71 | 0.63, 0.81 | 0.89 | 0.83, 0.96 |
| Other | 3.09 | 2.89, 3.31 | 0.63 | 0.53, 0.75 | 0.65 | 0.57, 0.74 |
| **Socioeconomic status of father** |  |  |  |  |  |  |
| Single/Unskilled/Other | Ref |  | NS |  | Ref |  |
| Partly Skilled | 0.87 | 0.78, 0.97 |  |  | 0.96 | 0.88, 1.04 |
| Manual | 0.83 | 0.75, 0.92 |  |  | 0.94 | 0.87, 1.01 |
| Non-Manual | 0.86 | 0.77, 0.97 |  |  | 0.86 | 0.78, 0.94 |
| Managerial and Technical | 0.93 | 0.83, 1.06 |  |  | 0.87 | 0.79, 0.95 |
| Professional | 0.81 | 0.69, 0.95 |  |  | 0.82 | 0.73, 0.93 |
| **Parental marital status** |  |  |  |  |  |  |
| Single | Ref |  | Ref |  | NS |  |
| Married | 0.55 | 0.51, 0.60 | 0.61 | 0.54, 0.68 |  |  |
| Separated/Divorced/Widowed | 0.78 | 0.66, 0.92 | 0.88 | 0.71, 1.10 |  |  |
| **Father's age at completion of education** |  |  |  |  |  |  |
| ≤15 year old | Ref |  | Ref |  | NS |  |
| 16-18 year old | 1.13 | 1.06, 1.22 | 1.24 | 1.13, 1.35 |  |  |
| ≥19 years old | 1.45 | 1.30, 1.62 | 1.44 | 1.27, 1.64 |  |  |
| **Parity (i.e. number of older siblings)** |  |  |  |  |  |  |
| 0 | Ref |  |  |  | Ref |  |
| 1 | 1.03 | 0.96, 1.10 |  |  | 1.09 | 1.03, 1.15 |
| 2 | 1.12 | 1.03, 1.22 |  |  | 1.10 | 1.03, 1.17 |
| 3 | 1.17 | 1.05, 1.30 |  |  | 1.12 | 1.03, 1.22 |
| >4 | 1.28 | 1.15, 1.42 |  |  | 1.19 | 1.10, 1.28 |
| **Method of contraception** |  |  |  |  |  |  |
| None | Ref |  | NS |  | Ref |  |
| Pill alone | 0.91 | 0.84, 0.98 |  |  | 0.91 | 0.86, 0.97 |
| Pill alone and other method | 0.90 | 0.75, 1.08 |  |  | 0.95 | 0.83, 1.10 |
| Other method | 0.80 | 0.74, 0.87 |  |  | 0.88 | 0.83, 0.94 |

**Table 3:** Estimated risk ratios and 95% confidence intervals for consistent predictors (selected in at least 50% of possible sweeps) of non-response at sweeps 4-9 (ages 26-46) in the 1970 British Cohort Study.

|  | Sweep 4 (age 26) |  | Sweep 5 (age 30) |  | Sweep 6 (age 34) |  | Sweep 7 (age 38) |  | Sweep 8 (age 42) |  | Sweep 9 (age 46) |  |
| --- | --- | --- | --- | --- | --- | --- | --- | --- | --- | --- | --- | --- |
|  | RR | 95% CI | RR | 95% CI | RR | 95% CI | RR | 95% CI | RR | 95% CI | RR | 95% CI |
| **Non-response at previous sweeps** |  |  |  |  |  |  |  |  |  |  |  |  |
| Incomplete vs complete response | 1.55 | 1.50, 1.61 | 3.58 | 3.31, 3.87 | 3.54 | 3.29, 3.81 | 3.36 | 3.09, 3.64 | 6.50 | 5.74, 7.35 | 4.55 | 4.14, 5.00 |
| **PREDICTORS from SWEEP 0 (age 0)** |  |  |  |  |  |  |  |  |  |  |  |  |
| **Sex** |  |  |  |  |  |  |  |  |  |  |  |  |
| Female vs Male | 0.78 | 0.75, 0.80 | 0.86 | 0.83, 0.90 | 0.90 | 0.87, 0.93 | 0.91 | 0.88, 0.94 | 0.91 | 0.88, 0.94 | NS | NS |
| **Socioeconomic status of father** |  |  |  |  |  |  |  |  |  |  |  |  |
| Single/Unskilled/Other | Ref |  | Ref |  | Ref |  | Ref |  | Ref |  | Ref |  |
| Partly Skilled | 0.95 | 0.90, 1.00 | 0.93 | 0.86, 1.01 | 0.95 | 0.90, 1.02 | 0.97 | 0.92, 1.03 | 0.95 | 0.89, 1.01 | 0.97 | 0.92, 1.03 |
| Manual | 0.90 | 0.85, 0.94 | 0.86 | 0.80, 0.92 | 0.92 | 0.87, 0.98 | 0.94 | 0.89, 0.99 | 0.90 | 0.85, 0.96 | 0.90 | 0.86, 0.95 |
| Non-Manual | 0.83 | 0.78, 0.89 | 0.87 | 0.79, 0.95 | 0.88 | 0.82, 0.95 | 0.89 | 0.83, 0.95 | 0.87 | 0.81, 0.94 | 0.88 | 0.82, 0.94 |
| Managerial and Technical | 0.82 | 0.76, 0.88 | 0.84 | 0.77, 0.92 | 0.86 | 0.79, 0.92 | 0.84 | 0.78, 0.90 | 0.79 | 0.73, 0.86 | 0.79 | 0.74, 0.85 |
| Professional | 0.83 | 0.75, 0.91 | 0.85 | 0.75, 0.96 | 0.87 | 0.78, 0.96 | 0.80 | 0.72, 0.88 | 0.79 | 0.70, 0.88 | 0.77 | 0.70, 0.85 |
| **Parity** |  |  |  |  |  |  |  |  |  |  |  |  |
| 0 | Ref |  | Ref |  | Ref |  | Ref |  | NS |  | Ref |  |
| 1 | 1.00 | 0.96, 1.05 | 0.93 | 0.88, 0.98 | 1.02 | 0.98, 1.07 | 0.99 | 0.95, 1.03 |  |  | 1.02 | 0.98, 1.06 |
| 2 | 1.04 | 0.99, 1.10 | 0.96 | 0.90, 1.03 | 1.09 | 1.04, 1.15 | 1.03 | 0.99, 1.08 |  |  | 1.06 | 1.01, 1.11 |
| 3 | 1.12 | 1.05, 1.19 | 0.97 | 0.89, 1.05 | 1.10 | 1.03, 1.18 | 1.12 | 1.06, 1.19 |  |  | 1.11 | 1.05, 1.17 |
| >4 | 1.16 | 1.10, 1.23 | 1.12 | 1.04, 1.21 | 1.18 | 1.11, 1.26 | 1.16 | 1.10, 1.23 |  |  | 1.12 | 1.06, 1.18 |
| **PREDICTORS from SWEEP 1 (age 5)** |  |  |  |  |  |  |  |  |  |  |  |  |
| **Household moves** |  |  |  |  |  |  |  |  |  |  |  |  |
| Per move | NS | NS | 1.04 | 1.02, 1.06 | 1.04 | 1.02, 1.05 | 1.04 | 1.02, 1.05 | NS |  | NS |  |
| **Copying Design Score** |  |  |  |  |  |  |  |  |  |  |  |  |
| Per unit | NS |  | NS |  | NS |  | 0.97 | 0.96, 0.98 | 0.97 | 0.96, 0.98 | 0.97 | 0.96, 0.99 |
| **PREDICTORS from SWEEP 2 (age 10)** |  |  |  |  |  |  |  |  |  |  |  |  |
| **British Ability Scales (BAS)** |  |  |  |  |  |  |  |  |  |  |  |  |
| Per unit | NS | NS | 0.99 | 0.98, 0.99 | NS | NS | 0.99 | 0.98, 0.99 | 0.97 | 0.96, 0.98 | 0.98 | 0.98, 0.99 |
| **PREDICTORS from**<br>**SWEEP 5 (age 30)** |  |  |  |  |  |  |  |  |  |  |  |  |
| **Voting in 1997 elections** |  |  |  |  |  |  |  |  |  |  |  |  |
| No (vs yes) | NA | NA | NA | NA | 1.14 | 1.08, 1.20 | 1.13 | 1.08, 1.17 | 1.11 | 1.05, 1.17 | 1.10 | 1.06, 1.14 |

### Restoring sample representativeness

In Figure 1, we show that the mean value of cognitive ability measured at age 5, among everybody who underwent all four tests (9529 participants) was 0.00 (95% CI: -0.03, 0.03), and among everybody who also remained alive and living in Great Britain by age 46 (sweep 9) was 0.01 (95% CI: -0.02, 0.04). We found that the mean value of cognitive ability at age 5 differed among respondents at sweep 9 [mean cognitive ability=0.12 (95% CI: 0.09, 0.16)], demonstrating substantial non-response bias. However, after MI, using all predictors of non-response at sweep 9 as auxiliary variables in the imputation model, we estimated a mean value of cognitive ability of 0.00 (95% CI: -0.03, 0.03), i.e. very close to the mean cognitive ability in our original sample. This suggests that the missing at random assumption was met and we were able to restore representativeness with respect to cognitive ability.

**Figure 1:**
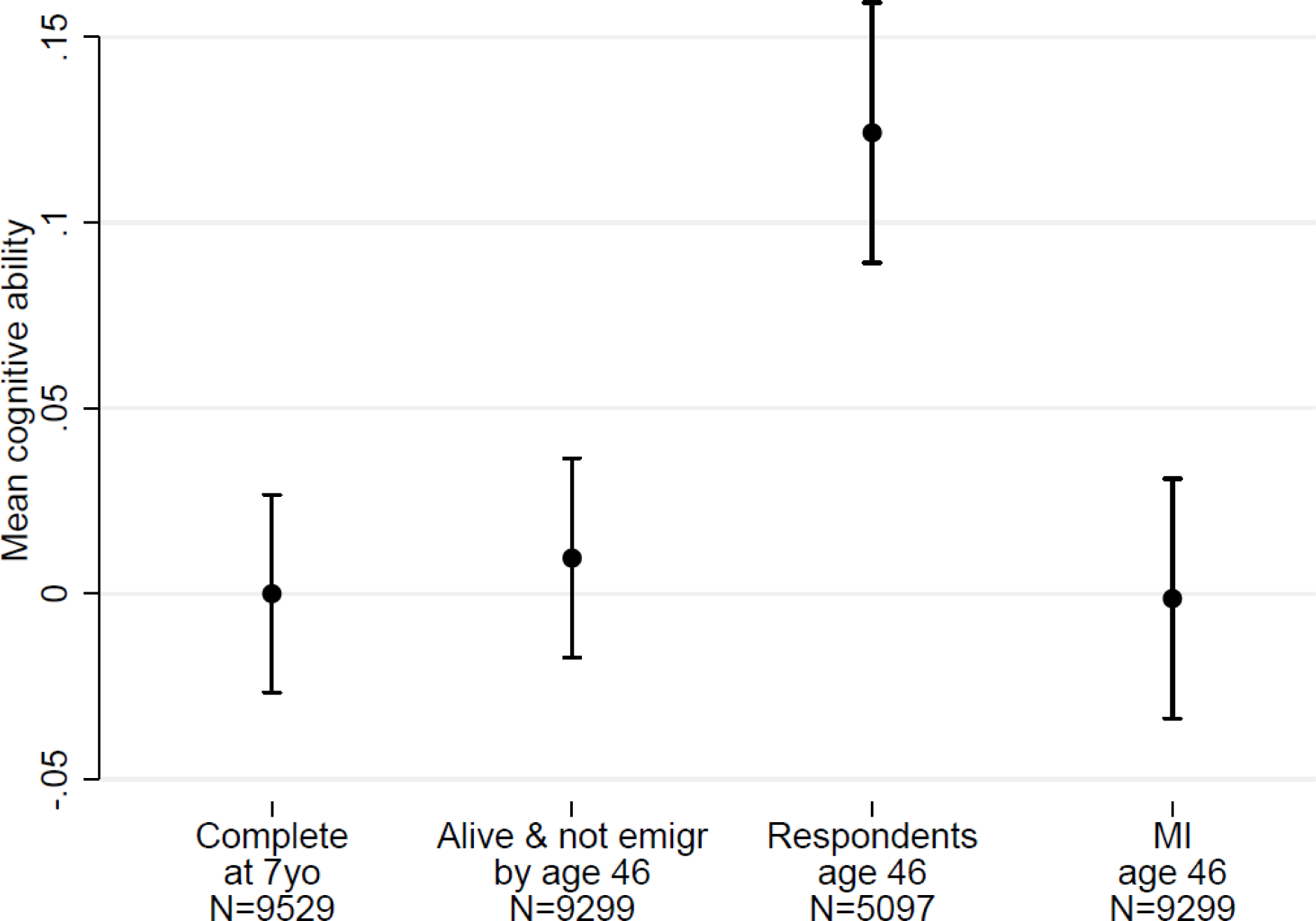
Internal validation through “traveling back in time”. Cognitive ability measured at age 5, estimated i) in the overall sample ii) among people who remained alive and did not emigrate by sweep 9 (age 46) iii) among respondents of sweep 9 (age 46) and iv) after performing multiple imputation to account for missing data in the non-respondents of sweep 9 (age 46)

In Figure 2, we observe in the 16,562 participants with complete data at birth that the percentage of fathers in the professional social class was 5.04% (95% CI: 4.71%, 5.37%), and among everybody who also remained alive and did not emigrate by age 46 (i.e. sweep 9, N=15,207) this was 5.00% (95% CI: 4.66%, 5.35%). We also found that this percentage differed [6.03% (95% CI: 5.51%, 6.55%)] among respondents at sweep 9 (N=7942), but after MI, we estimated the percentage to be 5.27% (95% CI: 4.76%, 5.78%), i.e. much closer to the percentage estimated among non-emigrants who remained alive.

**Figure 2:**
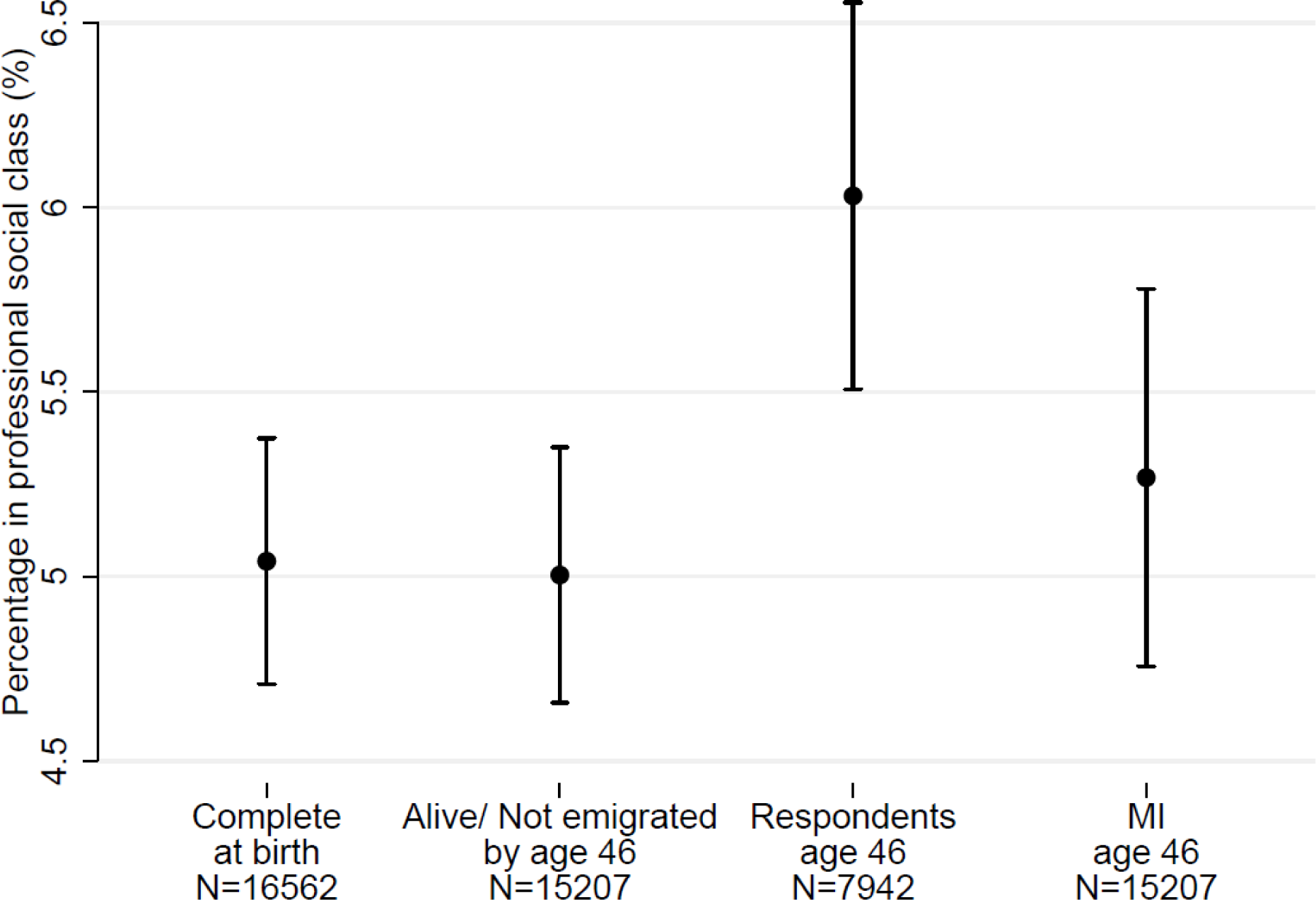
Internal validation through “traveling back in time”. Percentage of people with paternal professional work at sweep 0 estimated i) in the overall sample ii) among people who remained alive and did not emigrate by sweep 9 (age 46) iii) among respondents of sweep 9 (age 46) and iv) after performing multiple imputation to account for missing data in the non-respondents of sweep 9 (age 46)

Additionally, we examined the relationship between the cohort member’s father’s social class and the cohort member’s mortality after reaching the age of 26 while adjusting for sex and country of birth. In Figure 3, we present the hazard ratios (HRs) estimated among survivors at age 26 for the following comparisons: i) “partially skilled or manual” versus “unskilled/other father’s work” (HR=0.78 (95% CI: 0.48, 1.25)) ii) “non-manual or managerial” versus “unskilled/other” (HR=0.49 (95% CI: 0.29, 0.85)) and iii) “professional” versus “unskilled/other” (HR=0.84 (95% CI: 0.41, 1.72)). The results varied slightly when analysing respondents from sweep 4 (age 26) only. Specifically, the hazard ratios from the complete-case analysis were as follows: i) HR 0.60 (95% CI: 0.28, 1.27), ii) HR= 0.40 (95% CI: 0.17, 0.92) and iii) HR=1.00 (95% CI: 0.38, 2.61). Differences from the results using all available cohort members can again be interpreted as non-response bias. Subsequently, we performed MI using all predictors of non-response at sweep 4 as auxiliary variables in the imputation model. This allowed us to estimate hazard ratios that closely resembled those obtained from the initial sample. The hazard ratios after MI were as follows: i) HR=0.70 (95% CI: 0.38, 1.30), ii) HR= 0.52 (95% CI: 0.27, 1.00) and iii) HR=0.82 (95% CI: 0.35, 1.93).

**Figure 3:**
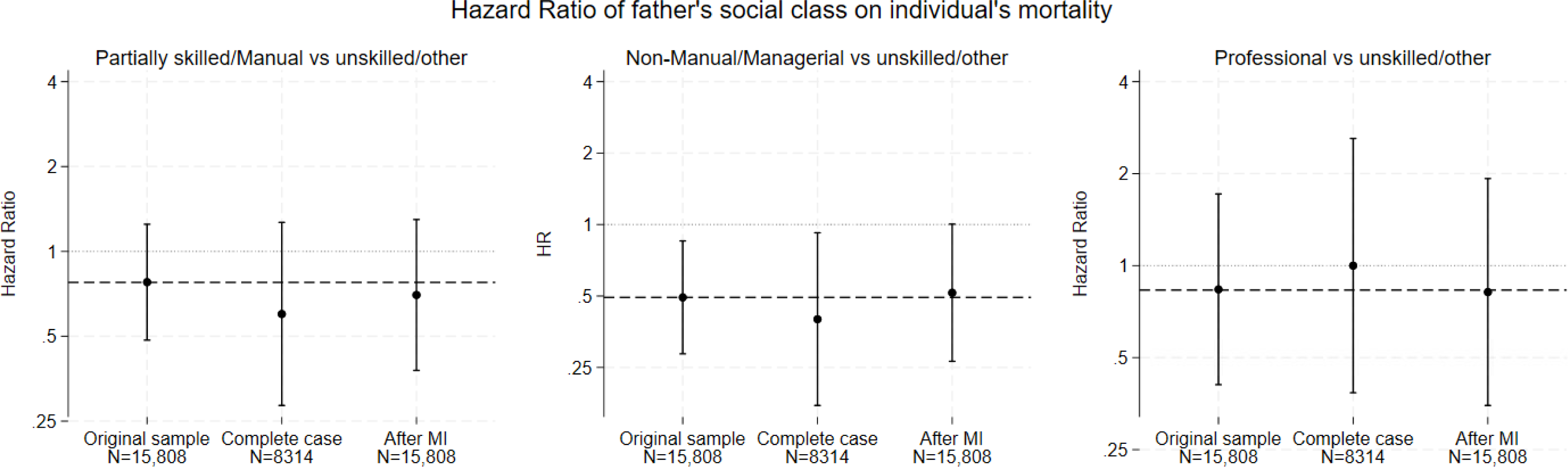
Internal validation through “traveling back in time”. Estimated hazard ratios of father’s social class on participants’ mortality after age 26 i) in the original sample of 15,842 individuals without missing data on the variable “father’s social class”, ii) among 8314 respondents (complete case) of sweep 4 (age 26) and iii) after multiple imputation to account for missing data on the non-respondents of sweep 4 (age 26)

The results for our BMI example at age 34, using data from HSE as a (potential) external benchmark, can be found in the appendix. In brief, after MI using all identified predictors of non-response at sweep 6 as auxiliary variables, the estimated BMI levels increased slightly to 26.63 kg/m^2^ (95% CI: 26.49, 26.76) for men and 25.35 kg/m^2^ (95% CI: 25.20, 25.50) for women (see Figure S2). These values were very close to the corresponding estimates from HSE and hence we implemented MI using delta adjustment, the delta values were relatively small.

## DISCUSSION

### 1. Summary of findings – comparison with the literature

We implemented a data driven approach to identify predictors of non-response in BCS70 that can be used as auxiliary variables for researchers to help address bias due to missing data. We built on our previous work in NCDS[1] and observed that the MAR assumption appeared plausible in different types of analysis in BCS70 once the rich data from earlier sweeps was incorporated into the analysis. We additionally illustrated how these predictors of non-response can be utilised when a MNAR mechanism is assumed more plausible, when there is information from an external source.

We harnessed information from 21,021 variables available in BCS70, aiming to provide a (relatively small) subset of variables that would be strongly related to non-response. We found associations between non-response at all 9 sweeps and predictors derived from data collected at previous sweeps. The results of this study are in line with our work in NCDS and with the literature on predictors of non-response in longitudinal cohorts. Specifically, we found that in the early sweeps (1-3; in which the parents of the participants were interviewed), the consistent predictors of non-response were parents’ non-participation in previous sweeps, participants’ country of birth, father’s disadvantaged socioeconomic status, greater number of older siblings for the cohort member, parents not being married and father’s higher age at completion of education. Our work from NCDS[1], as well as the corresponding literature on non-response in surveys, suggests that people from disadvantaged socio-economic backgrounds are less likely to respond[17,18]. Marital status[19] and ethnic background have also been found to be associated with non-response in surveys[20]. Moreover, we observed that in the later sweeps (4-9; in which the participants themselves are interviewed) the consistent predictors of non-response were being male, greater number of older siblings, disadvantaged paternal socioeconomic status, greater number of household moves, lower cognitive ability, and non-participation in the UK 1997 general elections and in the previous BCS70 sweeps. All these variables were identified in our previous work in NCDS and are consistent with the literature. The findings for non-participation in previous sweeps and for disadvantaged father’s socioeconomic status is consistent for both early sweeps (1-3) and later sweeps (4-9) and is in agreement with studies finding that disadvantaged socioeconomic background is linked to non-participation[1,17]. Moreover, it has been reported than women are more likely to respond to surveys, compared to men[18], increased moves increase the difficulty of making contact with participants[19,21], and early life cognitive ability was associated with survey participation[22], a finding potentially explained by the link between early life cognitive ability and educational attainment[23]. Additionally, greater number of older siblings[1] and disadvantaged paternal socioeconomic status[17] have also been documented as risk factors for non-participation. The identified predictors of non-response in BCS70 (from 7 to 16 variables for different sweeps) were fewer compared to NCDS (from 10 to 35 variables for different sweeps)[1], however qualitatively, the findings were very similar.

We have demonstrated that, by using the identified predictors of non-response from BCS70, we were able to restore sample representativeness for paternal social class at birth and cognitive ability collected at age 5, using data from only respondents at age 46. Additionally, the hazard ratios of paternal social class on adult mortality, when discarding information on non-respondents at age 26 and using MI, closely resembled those obtained from the complete sample. These findings suggest that for these example analyses, the MAR assumption can be considered plausible when incorporating observed predictors of non-response in principled methods of missing data handling. We further demonstrated how researchers can utilise these predictors of non-response and conduct analysis in cases where a valid external benchmark is available and they want to allow for the possibility of a MNAR mechanism,. In our example, we used HSE as a hypothetical external benchmark for BMI levels and used delta values after MI so that the mean BMI levels for men and women estimated from BCS70 would match the corresponding estimates from HSE. We will expand this work in other cohorts, including the National Survey of Health and Development[24,25] and the Millenium Cohort Study[26].

### 2. Strengths and limitations

We harnessed the rich information from 21,021 variables across 46 years of follow-up in BCS70 using a data driven approach to identify important predictors of non-response. Researchers usually address missing data problems from cohort studies using a-priori (theory-driven) approaches. If including auxiliary variables at all, they do not often select the best predictors of non-response using a validated procedure. Utilising a data-driven rather than a theory-driven approach allows us to identify the most important predictors of non-response for this particular context.

The main limitation of our study is that we used observational data, so the chance of measurement error or misclassification cannot be excluded. Moreover, the selection of the cut-offs of the p-values in our 3-stage process was arbitrary. The combination of these factors might have influenced the final selection of the predictors of non-response. However, there is evidence that our systematic missing data approach successfully restores sample representativeness and the expected distribution of a single variable or a relationship between two variables can be observed, when using the recommended auxiliary variables from our missing data strategy.

### 3. Conclusion

We implemented a systematic data-driven approach to identify predictors of non-response in BCS70. We provide a set of variables that researchers can utilise as auxiliary variables in their analyses, which can improve the plausibility of the MAR assumption and restore sample representativeness. Our approach can also be used when the MNAR assumption is more plausible, if a valid external benchmark is available.

## Data Availability

All data produced in the present study are available upon request to Dr Michail Katsoulis

