## Appendix for "A data driven approach to address missing data in the 1970 British birth cohort"

**Section 1: Description of the 3 stages and predictors of non-response**

In Figure S1 we present how we identified the predictors of non-response for each sweep. We followed a 3-stage analytic approach, starting from 21021 variables. We then excluded “routed” variables (questions that depend on a specific response to a previous question), excluded variables with >40% missing, excluded variables with categories including >98% of all cases, recoded variables with categories including <1% of cases if possible, and used index/score variables that combined information from many variables rather the individual constituent items wherever possible. We then used the remaining 1387 eligible variables as input. At each stage we modelled non-response with a Poisson model with robust standard errors (modified Poisson regression).

Below, we describe our 3-stage procedure for non-response at sweep t:

**Stage 1**: We used univariable modified Poisson regressions of non-response at sweep t on each potential predictor of non-response at sweep 0 up to sweep t –1 in complete case analysis. We kept predictors with p-value<0.001.

**Stage 2**: Multivariable modified Poisson regressions of non-response at sweep t on all predictors retained from stage 1, separately from sweep 0 up to sweep t –1. Keep predictors with p-value<0.05.

**Stage 3:** 4 variables (Sex, country of birth, participation in all previous sweeps and father’s socioeconomic status were used directly in Stage 3), were utilised directly in stage 3. These variables have been considered very strong predictors of non-response from our work in NCDS and from other survey studies. At this stage (stage 3), we performed multiple imputation (MI) using all retained variables plus non-response at sweep t in the imputation model. MI multivariable modified Poisson regressions for all retained predictors at sweep 0, up to sweep t–1, adjusted for predictors at all previous (but not subsequent) sweeps. We kept predictors with p-value <0.001.

The main difference from the analytic strategy documented for NCDS is that the approach for BCS70 was somewhat stricter (e.g. at Stage 1 the critical p-value was 0.001 versus 0.05 in NCDS, variables excluded from the list of potential predictors of non-response if they had missing data in >40% of the observations versus >50% in NCDS).

In Table S1, we present in detail the number of variables selected in stage 1, stage 2 and Stage 3 for predictors of non-response at each sweep from the previous sweeps and in Tables S2-S10, we present the results from all the predictors of non-response after stage 3, along with the estimated risk ratios and 95% confidence intervals.

Figure S1: Flowchart presenting the number of variables used to find the predictors of non-response

**Number of available variables: N=21021**

Excluded “routed” variables (questions that depend on a specific response to a previous question), variables with >40% missing, variables with categories including >98% of cases, recoded variables with categories <1%, excluded all binary variables with prevalence <1% of cases if possible, and used index/score variables that combined information from many variables rather the individual constituent items

**Variables to be used for Stage 1-3: N=967**

Application of the Stage 1-3 process

**Variables to be used after Stage 3:**

**Vary from N=7 (at sweep 1) to**

**N=16 (at sweep 9)**

|  | **NR 1** | **NR 2** | **NR 3** | **NR 4** | **NR 5** | **NR 6** | **NR 7** | **NR 8** | **NR9** |
| --- | --- | --- | --- | --- | --- | --- | --- | --- | --- |
| Wave 0  *Variables: 67* | S1: 28  S2: 15 | S1: 26  S2: 14 | S1: 30  S2: 13 | S1: 37  S2: 15 | S1: 30  S2: 14 | S1: 34  S2: 13 | S1: 35  S2: 13 | S1: 31  S2: 12 | S1: 35  S2: 11 |
| Wave 1  *Variables: 91* |  | S1: 15  S2: 8 | S1: 39  S2: 9 | S1: 56  S2: 6 | S1: 49  S2: 9 | S1: 55  S2: 11 | S1: 57  S2: 7 | S1: 49  S2: 8 | S1: 51  S2: 9 |
| Wave 2  *Variables: 162* |  |  | S1: 64  S2: 15 | S1: 88  S2: 14 | S1: 84  S2: 12 | S1: 91  S2: 13 | S1: 90  S2: 13 | S1: 82  S2: 9 | S1: 78  S2: 9 |
| Wave 3  *Variables: 108* |  |  |  | S1: 63  S2: 11 | S1: 39  S2: 7 | S1: 57  S2: 9 | S1: 65  S2: 12 | S1: 49  S2: 9 | S1: 54  S2: 7 |
| Wave 4  *Variables: 137* |  |  |  |  | S1: 35  S2: 7 | S1: 54  S2: 8 | S1: 64  S2: 13 | S1: 49  S2: 5 | S1: 53  S2: 10 |
| Wave 5  *Variables: 125* |  |  |  |  |  | S1: 56  S2: 11 | S1: 75  S2: 11 | S1: 56  S2: 12 | S1: 64  S2: 15 |
| Wave 6  *Variables: 148* |  |  |  |  |  |  | S1: 80  S2: 10 | S1: 69  S2: 17 | S1: 77  S2: 16 |
| Wave 7  *Variables: 51* |  |  |  |  |  |  |  | S1: 23  S2: 10 | S1: 27  S2: 8 |
| Wave 8  *Variables: 74* |  |  |  |  |  |  |  |  | S1: 39  S2: 5 |
| STAGE 3 without extra variables* | S3: 5  (out of 15 in total from S2) | S3: 7  (out of 22 in total from S2) | S3: 11  (out of 37 in total from S2) | S3: 10  (out of 46 in total from S2) | S3: 9  (out of 49 in total from S2) | S3: 12  (out of 65 in total from S2) | S3: 12  (out of 79 in total from S2) | S3: 9  (out of 82 in total from S2) | S3: 13  (out of 90 in total from S2) |
| Total with extra variables† | 7 | 9 | 14 | 13 | 12 | 15 | 15 | 14 | 16 |

Table S1: Number of variables selected in stage 1 (S1), stage 2 (S2) and Stage 3 (S3) for predictors of non-response at each sweep from the previous sweeps

*We do not include in these counts the variables sex, country of birth, participation in all previous sweeps and father’s socioeconomic status which were used directly in Stage 3

†We include in these counts the variables sex, country of birth, participation in all previous sweeps and father’s socioeconomic status

### **Table S2**. Estimated risk ratios and 95% confidence intervals for predictors of non-response at sweep 1 (age 7) (n = 17468).

| Sweep | Variable | RR | 95% CI |
| --- | --- | --- | --- |
| Sweep 0 | Marital Status |  |  |
| (age 0) | Single | 1.00 | (reference) |
|  | Married | 0.55 | 0.51, 060 |
|  | Divorced/Separated | 0.78 | 0.66, 0.92 |
|  | Parity |  |  |
|  | 0 | 1.00 | (reference) |
|  | 1 | 1.03 | 0.96, 1.10 |
|  | 2 | 1.12 | 1.03, 1.22 |
|  | 3 | 1.17 | 1.05, 1.30 |
|  | >4 | 1.28 | 1.15, 1.42 |
|  | Father’s social status |  |  |
|  | I Single, no work, unskilled other | 1.00 | (reference) |
|  | II partial work | 0.87 | 0.78, 0.97 |
|  | III manual work | 0.83 | 0.75, 0.92 |
|  | III non-manual work | 0.86 | 0.77, 0.97 |
|  | IV managerial/technical work | 0.93 | 0.83, 1.06 |
|  | V professorial work | 0.81 | 0.69, 0.95 |
|  | Country of Birth |  |  |
|  | England | 1.00 | (reference) |
|  | Wales | 0.69 | 0.59, 0.81 |
|  | Scotland | 1.13 | 1.04, 1.23 |
|  | Other | 3.09 | 2.89, 3.31 |
|  | Father’s age at completion of education |  |  |
|  | ≤15 year old | 1.00 | (reference) |
|  | 16-18 year old | 1.13 | 1.06, 1.22 |
|  | ≥19 years old | 1.45 | 1.30, 1.62 |
|  | Number of antenatal visits |  |  |
|  | Continuous (per visit) | 0.98 | 0.97, 0.98 |
|  | Method of contraception |  |  |
|  | None | 1.00 | (reference) |
|  | Pill alone | 0.91 | 0.84, 0.98 |
|  | Pill alone and other method | 0.90 | 0.75, 1.08 |
|  | Other method | 0.80 | 0.74, 0.87 |

### **Table S3**. Estimated risk ratios and 95% confidence intervals for predictors of non-response at sweep 2 (age 10) (n = 17447).

| Sweep | Variable | RR | 95% CI |
| --- | --- | --- | --- |
| Sweep 0 | Marital Status |  |  |
| (age 0) | Single | 1.00 | (reference) |
|  | Married | 0.61 | 0.54, 0.68 |
|  | Divorced/Separated | 0.88 | 0.71, 1.10 |
|  | Where was mother-date of birth |  |  |
|  | Consultant bed/ GP bed | 1.00 | (reference) |
|  | NHS unit | 0.81 | 0.72, 0.91 |
|  | Own home | 0.97 | 0.59, 1.59 |
|  | Other | 1.19 | 0.94, 1.09 |
|  | Country of Birth |  |  |
|  | England | 1.00 | (reference) |
|  | Wales | 0.65 | 0.53, 0.80 |
|  | Scotland | 0.71 | 0.63, 0.81 |
|  | Other | 0.63 | 0.53, 0.75 |
|  | Father’s age at completion of education |  |  |
|  | ≤15 year old | 1.00 | (reference) |
|  | 16-18 year old | 1.24 | 1.13, 1.35 |
|  | ≥19 years old | 1.44 | 1.27, 1.64 |
| Sweep 1 | Ethnic group (mother)  Non-european vs european | 1.55 | 1.31, 1.82 |
| (age 7) | Attitude to childhood independence  Per Unit increase in z-score | 0.92 | 0.88, 0.97 |
|  | Tenure of accommodation |  |  |
|  | Owned | 1.00 | (reference) |
|  | Bought | 0.85 | 0.73, 0.98 |
|  | Council rented | 0.79 | 0.68, 0.93 |
|  | Other | 1.07 | 0.91, 1.25 |
|  | Household moves (per move) | 1.10 | 1.06, 1.14 |
| Non response at  Previous sweeps  (i.e. sweep 1) |  | 4.53 | 4.20, 4.89 |

Results from sequential multiple imputation analyses in which potential predictors of non-response at a given sweep are adjusted for previously identified potential predictors of non-response at that sweep and previous sweeps (i.e. not at subsequent sweeps).

### **Table S4**. Estimated risk ratios and 95% confidence intervals for predictors of non-response at sweep 3 (age 16) (n = 17414).

| Sweep | Variable | RR | 95% CI |
| --- | --- | --- | --- |
| Sweep 0 | Father’s social status |  |  |
| (age 0) | I Single, no work, unskilled other | 1.00 | (reference) |
|  | II partial work | 0.96 | 0.88, 1.04 |
|  | III manual work | 0.94 | 0.87, 1.01 |
|  | III non-manual work | 0.86 | 0.78, 0.94 |
|  | IV managerial/technical work | 0.87 | 0.79, 0.95 |
|  | V professorial work | 0.82 | 0.73, 0.93 |
|  | Country of Birth |  |  |
|  | England | 1.00 | (reference) |
|  | Wales | 0.70 | 0.62, 0.79 |
|  | Scotland | 0.89 | 0.83, 0.96 |
|  | Other | 0.65 | 0.57, 0.74 |
|  | Age of mother at first birth |  |  |
|  | Per year | 0.99 | 0.98, 0.99 |
|  | Method of contraception |  |  |
|  | None | 1.00 | (reference) |
|  | Pill alone | 0.91 | 0.86, 0.97 |
|  | Pill & Other method | 0.95 | 0.83, 1.10 |
|  | Other Method | 0.88 | 0.83, 0.94 |
|  | Parity |  |  |
|  | 0 | 1.00 | (reference) |
|  | 1 | 1.09 | 1.03, 1.15 |
|  | 2 | 1.10 | 1.03, 1.17 |
|  | 3 | 1.12 | 1.03, 1.22 |
|  | >4 | 1.19 | 1.10, 1.28 |
|  | Certainty of last menstrual period |  |  |
|  | Certain vs uncertain | 1.10 | 1.05, 1.16 |
|  | Where was mother-date of birth |  |  |
|  | Consultant bed/ GP bed | 1.00 | (reference) |
|  | NHS unit | 0.84 | 0.79, 0.90 |
|  | Own home | 0.83 | 0.62, 1.11 |
|  | Other | 1.16 | 1.02, 1.32 |
| Sweep 1  (Age 6) | Type of accommodation |  |  |
|  | Detached | 1.00 | (reference) |
|  | Semi-detached | 0.95 | 0.88, 1.04 |
|  | Terrace | 1.02 | 0.93, 1.11 |
|  | Flat-Maisonette | 1.18 | 1.07, 1.30 |
|  | Rooms-other | 1.02 | 0.87, 1.20 |
|  | Ethnic group (father) |  |  |
|  | European UK | 1.00 | (reference) |
|  | European EU | 1.28 | 1.13, 1.45 |
|  | Other | 1.26 | 1.16, 1.37 |
|  | Neighbourhood group |  |  |
|  | Poor | 1.00 | (reference) |
|  | Average | 0.88 | 0.82, 0.95 |
|  | Well to do | 0.84 | 0.76, 0.93 |
|  | Rural | 0.78 | 0.70, 0.86 |
| Sweep 2  (Age 10) | Accommodation occupied by family |  |  |
|  | Flat self-contained | 1.00 | (reference) |
|  | House | 0.70 | 0.62, 0.79 |
|  | Other | 0.74 | 0.57, 0.95 |
|  | Estimated reading age (in years) |  |  |
|  | Continuous (per year) | 0.94 | 0.91, 0.96 |
|  | BAS Matrix - Total score |  |  |
|  | Continuous (per unit) | 0.98 | 0.98, 0.99 |
| Non response at  Previous sweeps  (i.e. sweeps 1 & 2) |  | 1.54 | 1.43, 1.66 |

Results from sequential multiple imputation analyses in which potential predictors of non-response at a given sweep are adjusted for previously identified potential predictors of non-response at that sweep and previous sweeps (i.e. not at subsequent sweeps).

### **Table S5**. Estimated risk ratios and 95% confidence intervals for predictors of non-response at sweep 4 (age 26) (n = 17287).

| Sweep | Variable | RR | 95% CI |
| --- | --- | --- | --- |
| Sweep 0 | Father’s social status |  |  |
| (age 0) | I Single, no work, unskilled other | 1.00 | (reference) |
|  | II partial work | 0.95 | 0.90, 1.00 |
|  | III manual work | 0.90 | 0.85, 0.94 |
|  | III non-manual work | 0.83 | 0.78, 0.89 |
|  | IV managerial/technical work | 0.82 | 0.76, 0.88 |
|  | V professorial work | 0.83 | 0.75, 0.91 |
|  | Age of mother at 1^st^ birth |  |  |
|  | Continuous (per year) | 0.99 | 0.98, 0.99 |
|  | Method of contraception |  |  |
|  | None | 1.00 | (reference) |
|  | Pill alone | 0.92 | 0.88, 0.96 |
|  | Pill & Other method | 0.91 | 0.82, 1.02 |
|  | Other Method | 0.88 | 0.84, 0.91 |
|  | Parity (i.e. number of older siblings) |  |  |
|  | 0 | 1.00 | (reference) |
|  | 1 | 1.00 | 0.96, 1.05 |
|  | 2 | 1.04 | 0.99, 1.10 |
|  | 3 | 1.12 | 1.05, 1.19 |
|  | >4 | 1.16 | 1.10, 1.23 |
|  | Certainty of last menstrual period |  |  |
|  | Certain vs uncertain | 1.07 | 1.03, 1.12 |
|  | Sex |  |  |
|  | Female vs male | 0.78 | 0.75, 0.80 |
| Sweep 1  (Age 6) | External score |  |  |
|  |  | 1.03 | 1.02, 1.04 |
|  | Harris scoring method |  |  |
|  | Per unit increase | 0.98 | 0.98, 0.99 |
| Sweep 2  (Age 10) | Gross family income | 0.97 | 0.96, 0.99 |
|  | Teacher Rutter assessment | 1.02 | 1.01, 1.03 |
|  | Number of household accessories | 0.98 | 0.96, 0.99 |
| Sweep 3  (Age 16) | Satisfaction with teen’s school progress |  |  |
|  | Very satisfied | 1.00 | (reference) |
|  | Fairly satisfied | 1.10 | 1.04, 1.16 |
|  | Not satisfied | 1.20 | 1.11, 1.29 |
| Non response at  Previous sweeps  (i.e. sweeps 1, 2 & 3) |  | 1.55 | 1.50, 1.61 |

Results from sequential multiple imputation analyses in which potential predictors of non-response at a given sweep are adjusted for previously identified potential predictors of non-response at that sweep and previous sweeps (i.e. not at subsequent sweeps).

### **Table S6**. Estimated risk ratios and 95% confidence intervals for predictors of non-response at sweep 5 (age 30) (n = 17035).

| Sweep | Variable | RR | 95% CI |
| --- | --- | --- | --- |
| Sweep 0 | Father’s social status |  |  |
| (age 0) | I Single, no work, unskilled other | 1.00 | (reference) |
|  | II partial work | 0.93 | 0.86, 1.01 |
|  | III manual work | 0.86 | 0.80, 0.92 |
|  | III non-manual work | 0.87 | 0.79, 0.95 |
|  | IV managerial/technical work | 0.84 | 0.77, 0.92 |
|  | V professorial work | 0.85 | 0.75, 0.96 |
|  | Sex |  |  |
|  | Female vs male | 0.86 | 0.83, 0.90 |
|  | Number of antenatal Visits |  |  |
|  | Per visit | 0.99 | 0.98, 0.99 |
|  | Parity (i.e. number of older siblings) |  |  |
|  | 0 | 1.00 | (reference) |
|  | 1 | 0.93 | 0.88, 0.98 |
|  | 2 | 0.96 | 0.90, 1.03 |
|  | 3 | 0.97 | 0.89, 1.05 |
|  | >4 | 1.12 | 1.04, 1.21 |
| Sweep 1  (Age 6) | Ethnic group (mother)  Non-european vs european | 1.18 | 1.07, 1.29 |
|  | Household moves |  |  |
|  | Per move | 1.04 | 1.02, 1.06 |
|  | Neighbourhood group |  |  |
|  | Poor | 1.00 | (reference) |
|  | Average | 0.93 | 0.87, 1.02 |
|  | Well to do | 0.81 | 0.73, 0.89 |
|  | Rural | 0.80 | 0.71, 0.89 |
| Sweep 2  (Age 10) | Score BAS Matrices | 0.99 | 0.98, 0.99 |
| Sweep 4  (Age 26) | How easy would you quit a job, if there  was no other job to go to (range: 1-5) |  |  |
|  | Continuous (per unit) | 0.94 | 0.91, 0.97 |
|  | N of accidents elsewhere |  |  |
|  | 0 | 1.00 | (reference) |
|  | 1 | 1.01 | 0.91, 1.12 |
|  | ≥2 | 1.34 | 1.14, 1.57 |
|  | Tenure of current address |  |  |
|  | Own/ Parents (rent-free) | 1.00 | (reference) |
|  | Buying on morgage | 0.75 | 0.65, 0.87 |
|  | Rent | 1.03 | 0.92, 1.17 |
|  | Other | 1.04 | 0.88, 1.24 |
| Non response at  Previous sweeps  (i.e. sweeps 1-4) |  | 3.58 | 3.31, 3.87 |

Results from sequential multiple imputation analyses in which potential predictors of non-response at a given sweep are adjusted for previously identified potential predictors of non-response at that sweep and previous sweeps (i.e. not at subsequent sweeps).

### **Table S7**. Estimated risk ratios and 95% confidence intervals for predictors of non-response at sweep 6 (age 30) (n = 16785).

| Sweep | Variable | RR | 95% CI |
| --- | --- | --- | --- |
| Sweep 0 | Father’s social status |  |  |
| (age 0) | I Single, no work, unskilled other | 1.00 | (reference) |
|  | II partial work | 0.95 | 0.90, 1.02 |
|  | III manual work | 0.92 | 0.87, 0.98 |
|  | III non-manual work | 0.88 | 0.82, 0.95 |
|  | IV managerial/technical work | 0.86 | 0.79, 0.92 |
|  | V professorial work | 0.87 | 0.78, 0.96 |
|  | Sex |  |  |
|  | Female vs male | 0.90 | 0.87, 0.93 |
|  | Ever a teenager mother |  |  |
|  | 0-19 | 1.00 | (reference) |
|  | 20 plus | 0.91 | 0.88, 0.95 |
|  | Parity (i.e. number of older siblings) |  |  |
|  | 0 | 1.00 | (reference) |
|  | 1 | 1.02 | 0.98, 1.07 |
|  | 2 | 1.09 | 1.04, 1.15 |
|  | 3 | 1.10 | 1.03, 1.18 |
|  | >4 | 1.18 | 1.11, 1.26 |
|  | Present marital status |  |  |
|  | Single | 1.00 | (reference) |
|  | Married | 0.81 | 0.76, 0.85 |
|  | Widowed/Divorced/Separated | 0.90 | 0.80, 1.00 |
|  | Method of contraception |  |  |
|  | None | 1.00 | (reference) |
|  | Pill alone | 0.94 | 0.90, 0.99 |
|  | Pill & Other method | 0.89 | 0.79, 1.01 |
|  | Other Method | 0.86 | 0.82, 0.90 |
| Sweep 1  (Age 6) | Household moves  Per move | 1.04 | 1.02, 1.05 |
|  | Number of household accessories |  |  |
|  | Per accessory | 0.96 | 0.95, 0.98 |
| Sweep 2  (Age 10) | Maths Test Score  Per mark | 0.99 | 0.99, 0.99 |
| Sweep 3  (Age 16) | Maths 0 level or equivalent  No vs yes | 1.17 | 1.07, 1.27 |
| Sweep 4  (Age 26) | Satisfaction about life (1-3) |  |  |
|  | per unit increase | 0.94 | 0.90, 0.97 |
| Sweep 5  (Age 30) | Does participant intend to move?  No vs Yes | 0.86 | 0.82, 0.90 |
|  | Did you vote in the 97 elections?  No vs Yes | 1.14 | 1.08, 1.20 |
|  | Do you like the area? (Units 1-5)  Per unit | 1.04 | 1.02, 1.06 |
| Non response at  Previous sweeps  (i.e. sweeps 1-4) |  | 3.54 | 3.29, 3.81 |

### **Table S8**. Estimated risk ratios and 95% confidence intervals for predictors of non-response at sweep 7 (age 36) (n = 16699).

| Sweep | Variable | RR | 95% CI |
| --- | --- | --- | --- |
| Sweep 0 | Father’s social status |  |  |
| (age 0) | I Single, no work, unskilled other | 1.00 | (reference) |
|  | II partial work | 0.97 | 0.92, 1.03 |
|  | III manual work | 0.94 | 0.89, 0.99 |
|  | III non-manual work | 0.89 | 0.83, 0.95 |
|  | IV managerial/technical work | 0.84 | 0.78, 0.90 |
|  | V professorial work | 0.80 | 0.72, 0.88 |
|  | Sex |  |  |
|  | Female vs male | 0.91 | 0.88, 0.94 |
|  | N of antenatal visits |  |  |
|  | Continuous (per visit) | 0.99 | 0.99, 1.00 |
|  | Parity (i.e. number of older siblings) |  |  |
|  | 0 | 1.00 | (reference) |
|  | 1 | 0.99 | 0.95, 1.03 |
|  | 2 | 1.03 | 0.99, 1.08 |
|  | 3 | 1.12 | 1.06, 1.19 |
|  | >4 | 1.16 | 1.10, 1.23 |
|  | Certainty of Data of last Menstrual Period |  |  |
|  | Certain vs uncertain | 1.12 | 1.08, 1.16 |
|  | Method of contraception |  |  |
|  | None | 1.00 | (reference) |
|  | Pill alone | 0.94 | 0.90, 0.98 |
|  | Pill & Other method | 0.90 | 0.81, 1.00 |
|  | Other method | 0.90 | 0.86, 0.94 |
| Sweep 1  (Age 6) | Household moves |  |  |
|  | Per move | 1.04 | 1.02, 1.05 |
|  | Copying design score |  |  |
|  | Per unit | 0.97 | 0.96, 0.98 |
| Sweep 2  (Age 10) | Maths Test Score  Continuous (Per mark) | 0.99 | 0.99, 1.00 |
|  | BASMATRIX  Continuous (Per unit) | 0.99 | 0.98, 0.99 |
| Sweep 4  (Age 26) | Age left education |  |  |
|  | 12-16 | 1.00 | (reference) |
|  | 17-18 | 0.88 | 0.80, 0.96 |
|  | 19 plus | 1.44 | 1.37, 1.52 |
| Sweep 5  (Age 30) | Does participant intend to have more children |  |  |
|  | Yes | 1.00 | (reference) |
|  | No | 1.13 | 1.08, 1.18 |
|  | Don’t know | 1.09 | 1.04, 1.15 |
|  | Financial situation |  |  |
|  | Living comfortably | 1.00 | (reference) |
|  | Doing alright | 0.97 | 0.92, 1.02 |
|  | Just about getting by | 1.00 | 0.95, 1.06 |
|  | Finding it quite difficult | 1.13 | 1.06, 1.21 |
|  | Finding it very difficult | 1.08 | 0.98, 1.19 |
|  | Voted in general elections 1997? |  |  |
|  | No vs yes | 1.13 | 1.08, 1.17 |
| Non response at  Previous sweeps  (i.e. sweeps 1-4) |  | 3.36 | 3.09, 3.64 |

Results from sequential multiple imputation analyses in which potential predictors of non-response at a given sweep are adjusted for previously identified potential predictors of non-response at that sweep and previous sweeps (i.e. not at subsequent sweeps).

### **Table S9**. Estimated risk ratios and 95% confidence intervals for predictors of non-response at sweep 8 (age 42) (n = 16638).

| Sweep | Variable | RR | 95% CI |
| --- | --- | --- | --- |
| Sweep 0 | Father’s social status |  |  |
| (age 0) | I Single, no work, unskilled other | 1.00 | (reference) |
|  | II partial work | 0.95 | 0.89, 1.01 |
|  | III manual work | 0.90 | 0.85, 0.96 |
|  | III non-manual work | 0.87 | 0.81, 0.94 |
|  | IV managerial/technical work | 0.79 | 0.73, 0.86 |
|  | V professorial work | 0.79 | 0.70, 0.88 |
|  | Country of Birth |  |  |
|  | England | 1.00 | (reference) |
|  | Wales | 1.01 | 0.94, 1.10 |
|  | Scotland | 1.14 | 1.08, 1.20 |
|  | Other | 0.98 | 0.88, 1.08 |
|  | Sex |  |  |
|  | Female vs male | 0.91 | 0.88, 0.94 |
|  | N of antenatal visits |  |  |
|  | Continuous (per visit) | 0.99 | 0.99, 1.00 |
|  | Certainty of Data of last Menstrual Period |  |  |
|  | Certain vs uncertain | 1.09 | 1.05, 1.14 |
| Sweep 1  (Age 6) | Copying design score  Continuous (Per unit) | 0.97 | 0.96, 0.98 |
| Sweep 2  (Age 10) | Score BAS Matrices  Per mark | 0.98 | 0.98, 0.99 |
| Sweep 4  (Age 26) | Net pay |  |  |
|  | Continuous – per pound | 1.00 | 1.00, 1.00 |
| Sweep 5  (Age 30) | Voted in general elections 1997?  No vs Yes | 1.11 | 1.05, 1.17 |
| Sweep 6  (Age 34) | Work overtime?  No vs Yes | 1.17 | 1.09, 1.25 |
|  | Should everyone behave responsibly?  No vs Yes | 1.26 | 1.12, 1.42 |
|  | How many times have you been found guilty in a criminal court?  ≥1 vs 0 | 0.56 | 0.46, 0.69 |
| Sweep 7  (Age 42) | Participant willing to be contacted for parents’ research project?  No vs yes | 1.30 | 1.19, 1.42 |
| Non response at  Previous sweeps  (i.e. sweeps 1-7) |  | 6.50 | 5.74, 7.35 |

Results from sequential multiple imputation analyses in which potential predictors of non-response at a given sweep are adjusted for previously identified potential predictors of non-response at that sweep and previous sweeps (i.e. not at subsequent sweeps).

### **Table S10**. Estimated risk ratios and 95% confidence intervals for predictors of non-response at sweep 9 (age 46) (n = 16585).

| Sweep | Variable | RR | 95% CI |
| --- | --- | --- | --- |
| Sweep 0 | Father’s social status |  |  |
| (age 0) | I Single, no work, unskilled other | 1.00 | (reference) |
|  | II partial work | 0.97 | 0.92, 1.03 |
|  | III manual work | 0.90 | 0.86, 0.95 |
|  | III non-manual work | 0.88 | 0.82, 0.94 |
|  | IV managerial/technical work | 0.79 | 0.74, 0.85 |
|  | V professorial work | 0.77 | 0.70, 0.85 |
|  | Country of Birth |  |  |
|  | England | 1.00 | (reference) |
|  | Wales | 1.04 | 0.98, 1.11 |
|  | Scotland | 1.12 | 1.08, 1.18 |
|  | Other | 0.93 | 0.85, 1.02 |
|  | Parity (i.e. number of older siblings) |  |  |
|  | 0 | 1.00 | (reference) |
|  | 1 | 1.02 | 0.98, 1.06 |
|  | 2 | 1.06 | 1.01, 1.11 |
|  | 3 | 1.11 | 1.05, 1.17 |
|  | >4 | 1.12 | 1.06, 1.18 |
|  | Certainty of last menstrual period |  |  |
|  | Certain vs uncertain | 1.08 | 1.04, 1.12 |
|  | Was lactation attempted |  |  |
|  | Not attempted vs attempted | 1.06 | 1.03, 1.10 |
|  | Number of antenatal visits |  |  |
|  | Per visit | 0.99 | 0.99, 1.00 |
| Sweep 1  (Age 6) | Copying designs score  Continuous (Per unit) | 0.97 | 0.96, 0.99 |
| Sweep 2  (Age 10) | Score BAS Matrices  Continuous (Per unit) | 0.98 | 0.98, 0.99 |
|  | Accommodation |  |  |
|  | Owned | 1.00 | (reference) |
|  | Bought | 0.95 | 0.89, 1.00 |
|  | Council rented | 1.04 | 0.98, 1.10 |
|  | Other rented | 1.01 | 0.92, 1.12 |
|  | Tied to occupation | 0.95 | 0.84, 1.07 |
| Sweep 5  (Age 30) | Had eczema or skin problems?  No vs yes | 1.10 | 1.04, 1.15 |
|  | Voted in general elections 1997? |  |  |
|  | No vs yes | 1.10 | 1.06, 1.14 |
| Sweep 6  (Age 36) | Is this address participant’s residence?  No vs yes | 1.11 | 1.05, 1.18 |
| Sweep 7  (Age 36) | Willing to be contacted for Parents Research Project  No vs Yes | 1.21 | 1.15, 1.28 |
|  | Any children aged 0-6 |  |  |
|  | No vs Yes | 1.11 | 1.05, 1.18 |
| Sweep 8  (Age 42) | Total score |  |  |
|  | Per mark | 0.98 | 0.97, 0.99 |
| Non response at  Previous sweeps  (i.e. sweeps 1-8) |  | 4.55 | 4.14, 5.00 |

Results from sequential multiple imputation analyses in which potential predictors of non-response at a given sweep are adjusted for previously identified potential predictors of non-response at that sweep and previous sweeps (i.e. not at subsequent sweeps).

**Section 2: MNAR analysis illustration with a (hypothetically valid) external benchmark.**

In this section we present an example of how our approach could be used if the missing data generating mechanism is suspected to be MNAR in the presence of an appropriate external benchmark. We conducted analysis on the BMI levels of individuals aged 34 from sweep 6 (age 34) in 2004. Multiple imputation with delta adjustment was employed, incorporating external information from the Health Survey for England (HSE)[1] as a hypothetically valid external benchmark. We note that we use data from HSE for illustrative purposes only, as it is challenging to be considered a valid external benchmark for BCS70. This is due to the fact that the sampling design of HSE was not aimed at those born in 1970, and it’s therefore not representative of the target population of BCS70.

Initially, we computed the mean BMI levels in BCS70 separately for men and women born in England from a complete-case analysis. Subsequently, we employed multiple imputation, utilizing all predictors related to non-response at sweep 6 as auxiliary variables, along with observed BMI from all the other sweeps. Finally, we applied multiple imputation with delta adjustment [2,3], ensuring that the estimated BMI levels from the HSE matched the mean BMI levels for men and women from the BCS70 dataset. The magnitude of the delta values also serves as an indicator of the plausibility of the MAR assumption: smaller delta values suggest the plausibility of MAR, while larger delta values favoured the MNAR assumption.

Specifically, we wanted to estimate the mean BMI levels for men and women in 2004, using BCS70 (sweep 6 - age 34). We calculated the mean BMI levels from an external source, Health Survey for England (HSE), and we wanted to observe if the estimates from BCS70, after implementing our missing data strategy would be close.

To estimate the mean BMI levels from HSE, separately for men and women at age 34, we assumed that the mean levels would be a weighted average of the estimates of (men and women) aged 25-34 and 35-44 (this was the available granularity from HSE). More specifically, we estimated

mean(BMI at 34)= 0.6* mean(BMI aged 25-34) + 0.4* mean(BMI aged 35-44)

For the standard error of the above estimate we followed the following procedure. We knew only the standard errors $\mathrm{se}\left( \mathrm{BMI}_{men,25-34} \right)$ and $\mathrm{se}\left( \mathrm{BMI}_{men,35-44} \right)$ for men and women aged 25-34 and 35-44, so we first calculated the variance for BMI for men and women aged 25-34 and 35-44. For men, we calculated:

$$Var\left( \mathrm{BMI}_{men,25-34} \right)={\mathrm{se}\left( \mathrm{BMI}_{men,25-34} \right)}^{2}*n_{men, 25-34}$$

$$Var\left( \mathrm{BMI}_{men,35-44} \right)={\mathrm{se}\left( \mathrm{BMI}_{men,35-44} \right)}^{2}*n_{men, 35-44}$$

Then we estimated the standard error for men aged 34, $\mathrm{se}\left( \mathrm{BMI}_{men,34} \right)$, from the formula

$$\mathrm{se}\left( \mathrm{BMI}_{men,34} \right)=\sqrt{{0.6}^{2}*\frac{Var\left( \mathrm{BMI}_{men,25-34} \right)}{n_{men, 25-34}}+{0.4}^{2}*\frac{Var\left( \mathrm{BMI}_{men,35-44} \right)}{n_{men, 35-44}}}$$

We then performed the same procedure for women as well to estimate $\mathrm{se}\left( \mathrm{BMI}_{women,34} \right)$

To investigate the plausibility of the MNAR assumption compared to the MAR assumption when calculating mean BMI levels, we conducted an analysis on the BMI levels of individuals aged 34 from sweep 6 (age 34) in 2004. Multiple imputation with delta adjustment was employed, incorporating information from HSE as an external benchmark.

The delta values were relatively small (0.53kg/m^2^ in men and 1.77 kg/m^2^ in women), which is an indication that t the MAR assumption was plausible, see Figure S2 below.

**Results**

The mean BMI levels from HSE were 26.89 kg/m^2^ (95% CI: 26.59, 27.19) for men and 26.17 kg/m^2^ (95% CI: 25.81, 26.52) for women (Figure S2, Appendix). The mean BMI levels from BCS70 from the complete case analysis of participants born in England were 26.51 kg/m^2^ (95% CI: 26.37, 26.65) for men and 25.12 kg/m^2^ (95% CI: 24.96, 25.29) for women, lower than those in HSE. If the HSE data are taken to be population-representative, this suggests that BCS70 sweep 6 respondents had lower BMI than would be expected (i.e. bias due to non-response). When we implemented multiple imputation, using all identified predictors of non-response at sweep 6 as auxiliary variables, the estimated BMI levels increased slightly to 26.63 kg/m^2^ (95% CI: 26.49, 26.76) for men and 25.35 kg/m^2^ (95% CI: 25.20, 25.50) for women, though remained below the HSE levels. The delta values that would be needed if using delta adjustment would be relatively small (0.53 kg/m^2^ in men and 1.77 kg/m^2^ in women), which could have been an indication that the MAR assumption was plausible. However, HSE is not a valid external benchmark, so our ability to draw such conclusions is limited, even when the delta values are small.

Figure S2: Comparison of mean BMI levels in men (upper panel) and women (upper panel) at age 34 in 2004 estimated from Health Survey from England (HSE) and BCS 70. Multiple imputation using delta adjustment was used in BCS 70 to match mean estimates values of BCS 70 with HSE.

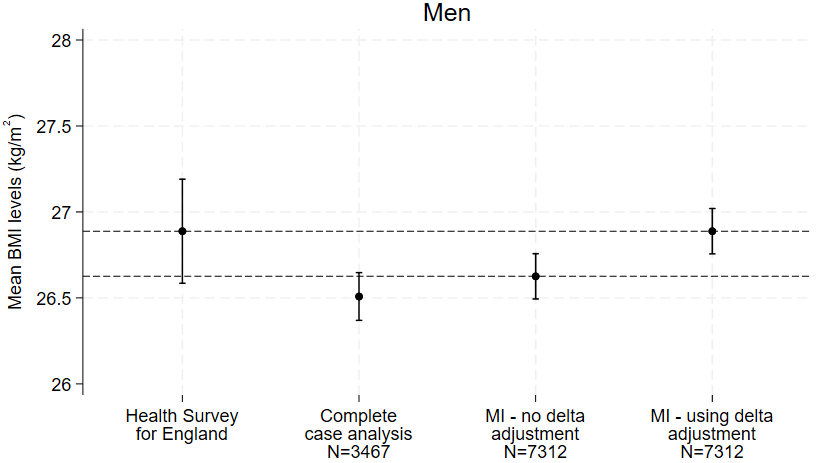

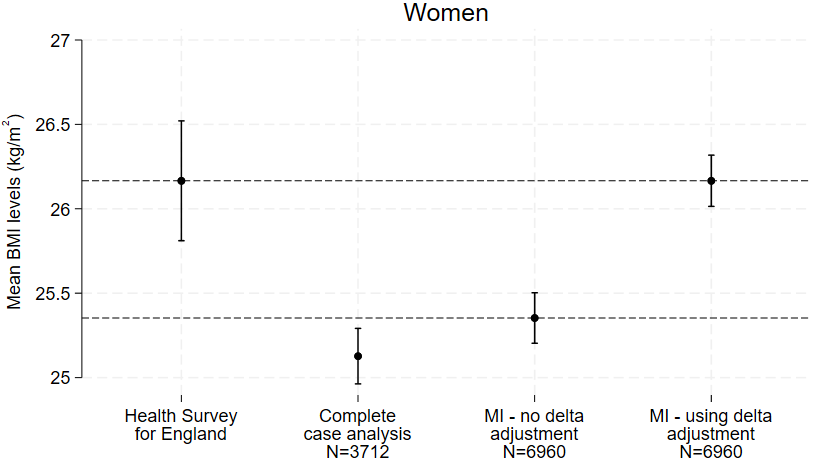
